# Post-hoc Estimation of a Quantitative Restriction Spectrum Imaging Biomarker for Prostate Cancer Detection Using Conventional MRI

**DOI:** 10.1101/2025.10.10.25337687

**Authors:** Deondre D Do, Christopher C Conlin, Aditya Bagrodia, Matthew Cooperberg, Michael E Hahn, Mukesh Harisinghani, Gary Hollenberg, Juan Javier-Desloges, Sophia C. Kamran, Christopher J Kane, Kang-Lung Lee, Michael A Liss, Daniel JA Margolis, Paul M Murphy, Nabih Nakrour, Michael A. Ohliger, Thomas Osinski, Rebecca Rakow-Penner, Mariluz Rojo Domingo, Amirali Salmasi, Ahmed S Shabaik, Yuze Song, Shaun Trecarten, Natasha Wehrli, Eric P Weinberg, Sean Woolen, Anders M Dale, Tyler M Seibert

## Abstract

**Background:** Multiparametric MRI is useful for early detection of clinically significant prostate cancer (csPCa), but its standard Apparent Diffusion Coefficient (ADC) has limited utility as a quantitative metric for automated, patient-level detection of csPCa. Restriction Spectrum Imaging (RSI), an advanced diffusion technique, yields a quantitative biomarker (RSIrs) that improves csPCa detection. RSIrs is typically calculated from a dedicated multi-*b*-value acquisition. RSIrs estimated from conventional MRI has not been studied.

**Purpose:** To evaluate the accuracy and validity of RSI metrics estimated *post-hoc* from conventional diffusion-weighted imaging (DWI) to serve as a viable surrogate for a dedicated RSI acquisition.

**Materials and Methods:** We conducted a retrospective, multi-center study of patients with both a dedicated RSI acquisition and conventional DWI. We compared three different RSI restriction score (RSIrs) calculation methods: from the dedicated acquisition (RSIrs_dedicated_), from conventional DWI alone (RSIrs_post-hoc_), and from a combination of conventional DWI with only the high *b*-values from the RSI acquisition (RSIrs_combo_). We compared these methods for quantitative agreement and csPCa detection performance (Area under the Receiver Operating Characteristic [AUC, 95% Confidence Interval]) of maximum RSIrs (RSIrs_max_) in the prostate compared to that of minimum ADC (ADC).

**Results:** Data from n=1095 patients (16 centers) were analyzed. *Post-hoc* RSIrs_max_ differed systematically from RSIrs_dedicated_ by a median of +156 (RSIrs_post-hoc_) and −59 (RSIrs_combo_), respectively. AUCs for csPCa detection were 0.51 [0.47,0.54], 0.60 [0.57,0.64], 0.70 [0.67,0.74], and 0.77 [0.74,0.80] for ADC, RSIrs_post-hoc_, RSIrs_combo_, and RSIrs_dedicated_, respectively.

**Conclusion:** Even when estimated using conventional DWI, RSIrs is a superior quantitative biomarker to ADC for automated, patient-level detection of csPCa. A dedicated RSI acquisition gives the best performance. A compromise would be to acquire high *b-*values (1500 s/mm^2^, 2500 s/mm^2^) to complement low *b-*values (<1000 s/mm^2^) from conventional DWI.

## Introduction

Multiparametric MRI (mpMRI) is useful for detection and management of prostate cancer [Ahmed et al. 2017, Kasivisvanathan et al. 2018, Moses et al. 2023, Mottet et al. 2021, Kerkmeijer et al. 2021, Menne et al. 2025]. mpMRI includes Diffusion-weighted imaging (DWI) that permits calculation of a quantitative metric, the Apparent Diffusion Coefficient (ADC). This metric is used to localize restricted water movement, a characteristic feature of hypercellular csPCa. Used in tandem with other image sequences in accordance with the Prostate Imaging Reporting & Data System (PI-RADS), ADC maps help detect csPCa lesions on prostate MRI [Turkbey et al. 2019]. However, the utility of ADC by itself as a quantitative metric is limited, in part because a lesion must be identified by an experienced radiologist.

The model for ADC oversimplifies the complex, multi-structural nature of water diffusion in human tissue [White et al. 2014, Zhong et al. 2023, Dornisch et al. 2024]. Restriction Spectrum Imaging (RSI) is a more sophisticated DWI technique that better accounts for tissue microstructure [Conlin et al. 2021]. The typical RSI framework models the diffusion MRI signal as a combination of exponential decays, corresponding to four distinct microcompartments: intracellular, extracellular, free diffusion (e.g., in the lumina of glands), and vascular flow. A quantitative biomarker derived from RSI—the RSI restriction score (RSIrs)—is useful for localizing csPCa. Previous work has demonstrated that RSIrs is superior to standard ADC for detecting csPCa on a patient-level, defining tumor boundaries for radiotherapy targeting, and helping non-experts identify csPCa on mpMRI [Zhong et al. 2023, Lui et al., 2023, Rojo Domingo et al., 2024, Rojo Domingo et al. 2024].

The RSI modeling framework can be applied to any conventional DWI dataset, provided enough non-zero *b*-values were acquired to estimate the model parameters. However, it is unclear whether RSIrs can be accurately estimated from conventional DWI data used for PI-RADS. Estimating RSI metrics with conventional DWI could be attractive for retrospective studies where only conventional DWI has been acquired. Additionally, some centers might want to implement RSI but prefer not to change their acquisition protocols.

We retrospectively applied the prostate RSI model to conventional DWI data in a cohort of patients who were scanned with both conventional DWI and a dedicated RSI sequence. This allowed for a direct, within-patient comparison of *post-hoc* RSIrs (RSIrs_post-hoc_) to dedicated RSIrs (RSIrs_dedicated_). We also investigated RSIrs estimated from a combination of conventional DWI and RSI data (RSIrs_combo_) to determine if supplementing conventional DWI with additional *b*-values could improve performance. We hypothesized that *post-hoc* RSI would have some quantitative agreement with RSI and would remain superior to conventional ADC for fully automated, patient-level detection of csPCa.

## Methods

### Study Population and Data Source

This retrospective analysis utilized a multi-center dataset drawn from six imaging centers within the Quantitative Prostate Imaging Consortium, including the University of California San Diego (UCSD) Center for Translational Imaging and Precision Medicine (CTIPM), University of California San Diego Health (UCSDH), University of California San Francisco (UCSF), Harvard Massachusetts General Hospital (MGH), University of Rochester Medical Center (URMC), and University of Texas Health Sciences Center San Antonio (UTHSCA). All data were collected under protocols approved by each center’s respective Institutional Review Board. In accordance with these protocols, some data were used after written informed consent as part of prospective studies, while others were analyzed under a waiver of consent for the retrospective use of clinical data (at centers using RSI clinically).

The study cohort comprised men aged 18 years or older who underwent 3T prostate MRI with RSI between January 2016 and March 2024 for the evaluation of suspected or known csPCa. This included patients with no prior diagnosis (i.e., due to elevated PSA). A composite reference standard was used to determine the ground truth for the presence of csPCa. Patient cases were considered non-csPCa if they had histopathologic confirmation (benign or Grade Group 1) from a biopsy performed within six months of the MRI. Given that biopsy is generally not performed for negative MRI, we also included as non-csPCa cases with PI-RADS 1-2 combined with a low PSA density (PSAD ≤ 0.15 ng/mL^2^). This introduces the possibility of a small number of cases with csPCa incorrectly assumed to be non-csPCa, which could artificially diminish the measured performance of RSI or other metrics. Though this risk is small, the approach confers better statistical power to measure discrimination, overall. Patients were excluded if they had received treatment for prostate cancer prior to their MRI or if they had metallic implants that could cause significant imaging artifacts. Patients must have had both an acquisition of a conventional DWI sequence and a separate, dedicated RSI sequence during the same imaging session.

### RSI Data Acquisition, Processing, and Modeling

All DWI data (conventional and RSI) underwent a standardized pre-processing pipeline that included correction for background noise, geometric distortion correction, gradient nonlinearities, *B*_*0*_ inhomogeneities and eddy currents [Holland et al. 2010, White et al. 2014]. All conventional DWI data were resampled into the same imaging space (determined by the dedicated RSI acquisition), followed by rigid-body registration performed using the *b*=0 (s/mm^2^) acquisitions of each available scan to correct for minor patient motion that may have occurred between the acquisitions. Automated whole-prostate segmentations were generated for each patient using an in-house deep-learning based tool [Song et al. 2024]. To account for signal intensity differences between the two scans, a scaling factor was calculated from the ratio of the median signal intensities within the prostate mask on the *b*=0 (s/mm^2^) images of the available data. The final *post-hoc* DWI volume is then processed using the multi-compartmental RSI model. For conventional DWI with inadequate (< 4) *b*-values to model the 4 compartment RSI model parameters, a simplified 3-compartment RSI model framework was used instead [Conlin et al. 2021]. Attempting to fit a 4 compartment RSI model with insufficient data points leads to unstable model solutions not representative of diffusion in the tissue microcompartments. The RSIrs biomarker is then derived from the intracellular signal component (RSI-C_1_), normalized by the median DWI signal at either *b* = 0 (s/mm^2^) of the prostate to reduce scanner-dependent signal variation (Equation 1). For protocols without a *b* = 0 (s/mm^2^) acquisition in their conventional DWI scans, normalization was done with the median DWI signal of the patient’s lowest *b*-value acquisition.

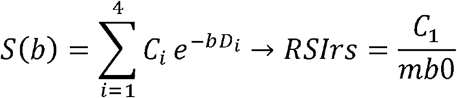

Equation 1: Formula to compute RSIrs where S(*b*) represents the RSI signal. C_i_ denotes the RSI compartment signal contribution, D_i_ is the compartmental diffusion coefficient, and mb0 is the median DWI signal in the prostate (x ∈ {0, 50})

For this study, four distinct quantitative RSIrs maps were generated for each patient:

1. RSIrs_dedicated_ (Reference Standard): Calculated with diffusion data from the complete dedicated multi-*b*-value RSI dataset.
2. RSIrs_post-hoc_: Estimated by applying the RSI model using only the *b*-values available from the patient’s conventional DWI scan (including a full FOV [Full FOV] and smaller FOV focused on the prostate [FOCUS])
3. RSIrs_combo_: Estimated from a combined dataset comprising the conventional DWI *b*-values (*b* ≤ 1000 s/mm^2^ from the Full FOV scan) supplemented with a subset of high *b*-values (> 1000 s/mm^2^) from the dedicated RSI scan.
4. ADC: Apparent Diffusion Coefficient maps were calculated in standard fashion using *b*-values <1000 s/mm^2^. To establish the baseline for a simple, automated quantitative biomarker, the minimum ADC value across the entire prostate gland was recorded for each patient. This whole-gland metric was chosen to test ADC’s capability of providing a single patient-level quantitative biomarker for detecting csPCa with the understanding that it is susceptible to known confounders like benign prostatic tissue. It is not how ADC is typically used clinically (which would require subjective identification of suspicious lesions by experienced radiologists). Rather, it is an analogous comparator to RSIrs as a quantitative biomarker for automated, patient-level detection.

All post-processing was performed using custom scripts developed in MATLAB (MathWorks, Natick, MA).

### Statistical Analysis

All statistical analyses were performed in Python (v3.8) using the scipy, pandas, and scikit-learn libraries. As the maximum RSIrs is typically used for detecting csPCa, we use this metric as our standard for comparison of *post-hoc* estimations of RSIrs to dedicated RSIrs. Our evaluation focused on two primary outcomes:

#### Quantitative Bias and Agreement

Bland-Altman analyses were performed to find the maximum RSIrs difference (bias) and 95% limits of agreement between the *post-hoc* RSIrs estimation methods and the dedicated RSI reference standard. We directly compared the *post-hoc* methods to each other by calculating the error in maximum RSIrs of each method to the reference standard for each patient. The statistical significance of the median bias was assessed using a two-sided Wilcoxon signed-rank test, with significance set at alpha 0.05.

#### csPCa Detection Performance

To compare the automated/quantitative, patient-level csPCa detection potential of each metric, we generated Receiver Operating Characteristic (ROC) curves and calculated the Area Under the Curve (AUC) for each of the four imaging metrics. 95% confidence intervals (95% CIs) were generated using 10,000 patient bootstraps for each metric. We conducted the above analyses stratifying by protocol, model framework (3 vs 4 compartment), and in a subset of patients with PI-RADS lesions contoured on MRI and tumors contoured on whole-mount histopathology by a subspecialist expert radiologist and pathologist, respectively. Co-registration of whole-mount histopathology to MRI was done using the previously validated RAPSODI software [Rusu et al. 2020].

## Results

1095 patients were eligible for this study (Figure 1; Table 1), MRI data were gathered from 16 MRI scanners (3 unique models) from 2 vendors (GE Healthcare, Waukesha, WI, USA; SIEMENS Healthineers, Erlangen, Germany) and 12 unique DWI protocols (6 conventional and 6 dedicated RSI). Detailed parameters can be found in Supplementary Table 1.

**Table 1.**
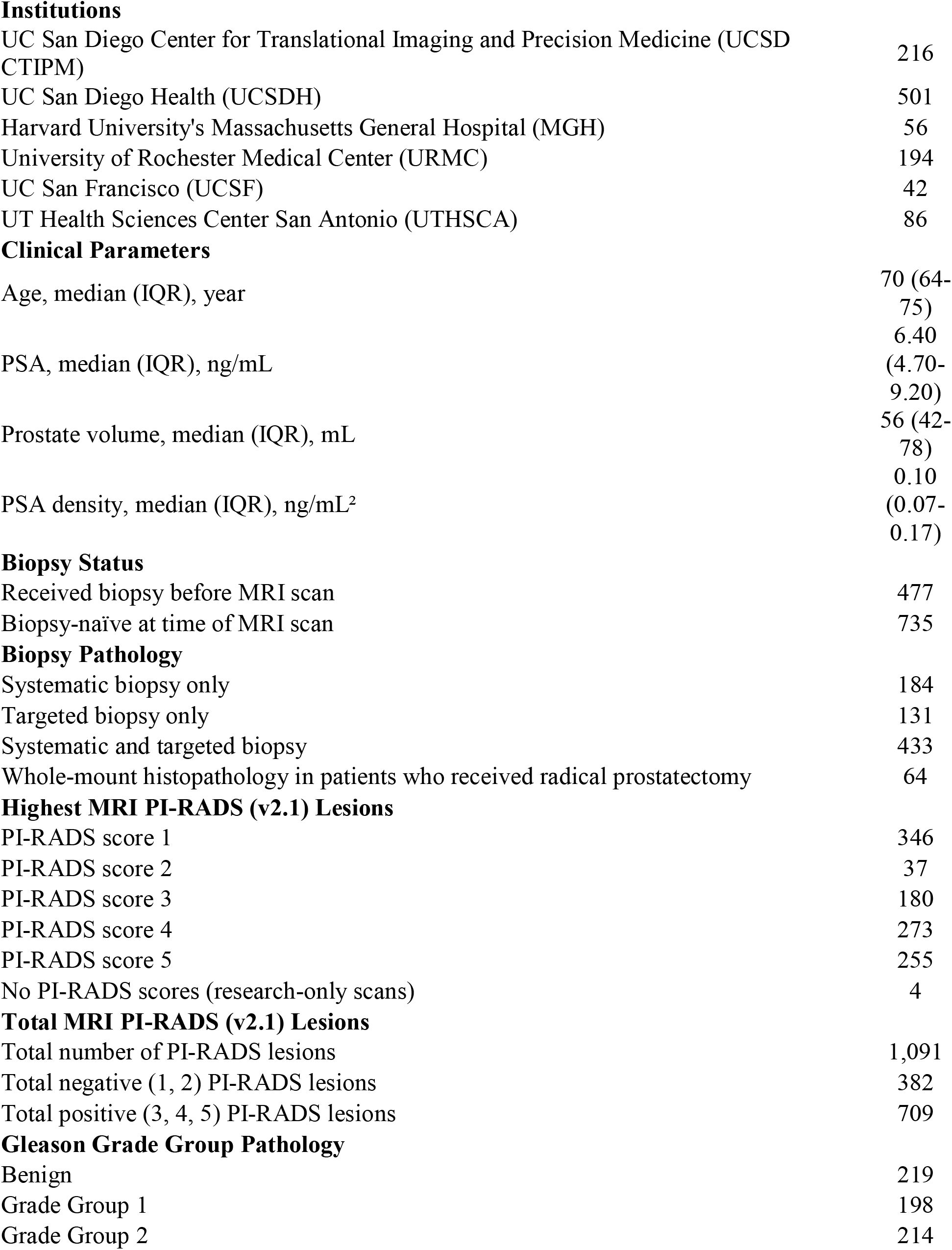
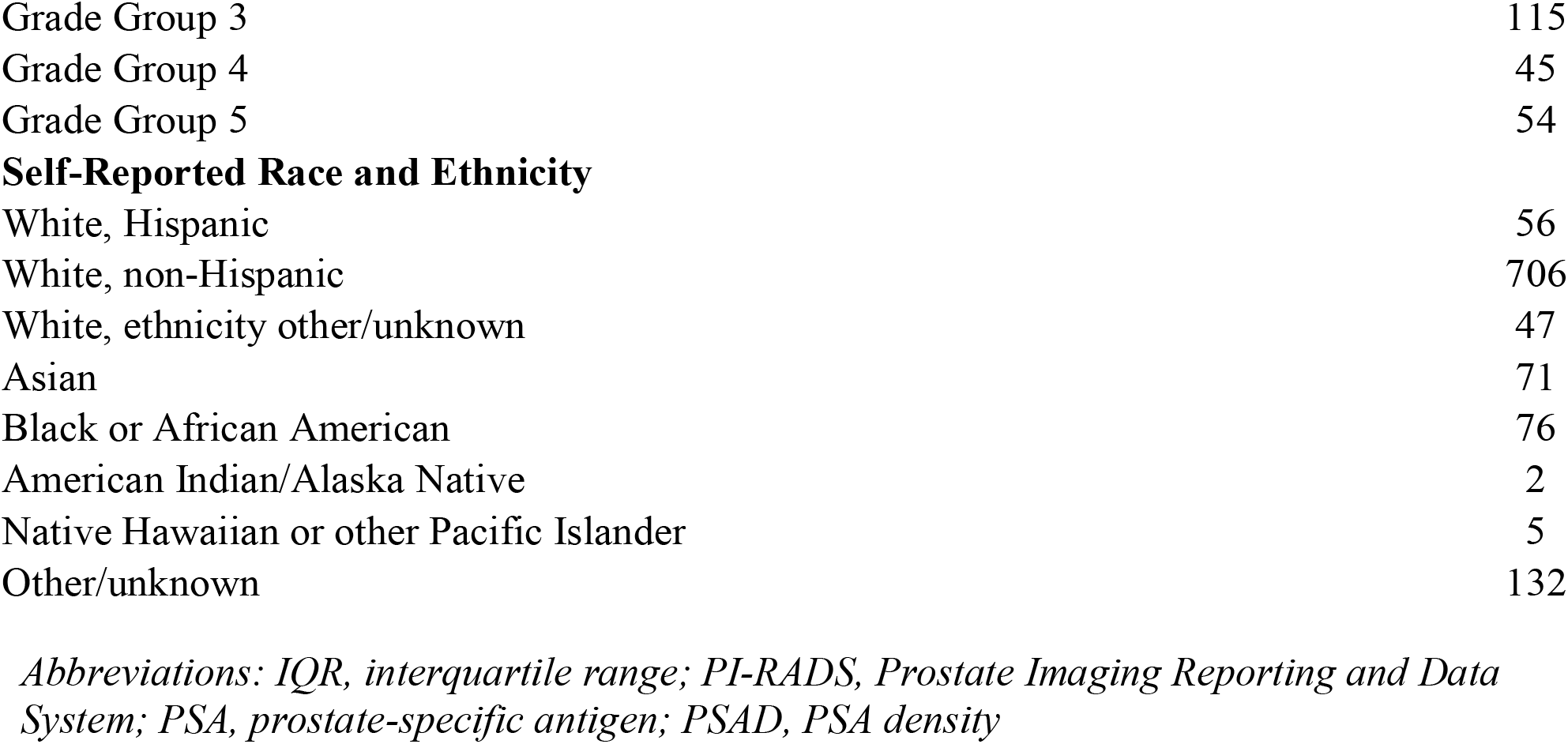
Patient Characteristics.

**Institutions**
|  |  |
| --- | --- |
| UC San Diego Center for Translational Imaging and Precision Medicine (UCSD CTIPM) | 216 |
| UC San Diego Health (UCSDH) | 501 |
| Harvard University's Massachusetts General Hospital (MGH) | 56 |
| University of Rochester Medical Center (URMC) | 194 |
| UC San Francisco (UCSF) | 42 |
| UT Health Sciences Center San Antonio (UTHSCA) | 86 |
| **Clinical Parameters** |  |
| Age, median (IQR), year | 70 (64-75) |
| PSA, median (IQR), ng/mL | 6.40 (4.70-9.20) |
| Prostate volume, median (IQR), mL | 56 (42-78) |
| PSA density, median (IQR), ng/mL<sup>2</sup> | 0.10 (0.07-0.17) |
| **Biopsy Status** |  |
| Received biopsy before MRI scan | 477 |
| Biopsy-naïve at time of MRI scan | 735 |
| **Biopsy Pathology** |  |
| Systematic biopsy only | 184 |
| Targeted biopsy only | 131 |
| Systematic and targeted biopsy | 433 |
| Whole-mount histopathology in patients who received radical prostatectomy | 64 |
| **Highest MRI PI-RADS (v2.1) Lesions** |  |
| PI-RADS score 1 | 346 |
| PI-RADS score 2 | 37 |
| PI-RADS score 3 | 180 |
| PI-RADS score 4 | 273 |
| PI-RADS score 5 | 255 |
| No PI-RADS scores (research-only scans) | 4 |
| **Total MRI PI-RADS (v2.1) Lesions** |  |
| Total number of PI-RADS lesions | 1,091 |
| Total negative (1, 2) PI-RADS lesions | 382 |
| Total positive (3, 4, 5) PI-RADS lesions | 709 |
| **Gleason Grade Group Pathology** |  |
| Benign | 219 |
| Grade Group 1 | 198 |
| Grade Group 2 | 214 |
| Grade Group 3 | 115 |
| Grade Group 4 | 45 |
| Grade Group 5 | 54 |
| **Self-Reported Race and Ethnicity** |  |
| White, Hispanic | 56 |
| White, non-Hispanic | 706 |
| White, ethnicity other/unknown | 47 |
| Asian | 71 |
| Black or African American | 76 |
| American Indian/Alaska Native | 2 |
| Native Hawaiian or other Pacific Islander | 5 |
| Other/unknown | 132 |

*Abbreviations: IQR, interquartile range; PI-RADS, Prostate Imaging Reporting and Data System; PSA, prostate-specific antigen; PSAD, PSA density*

**Figure 1:**
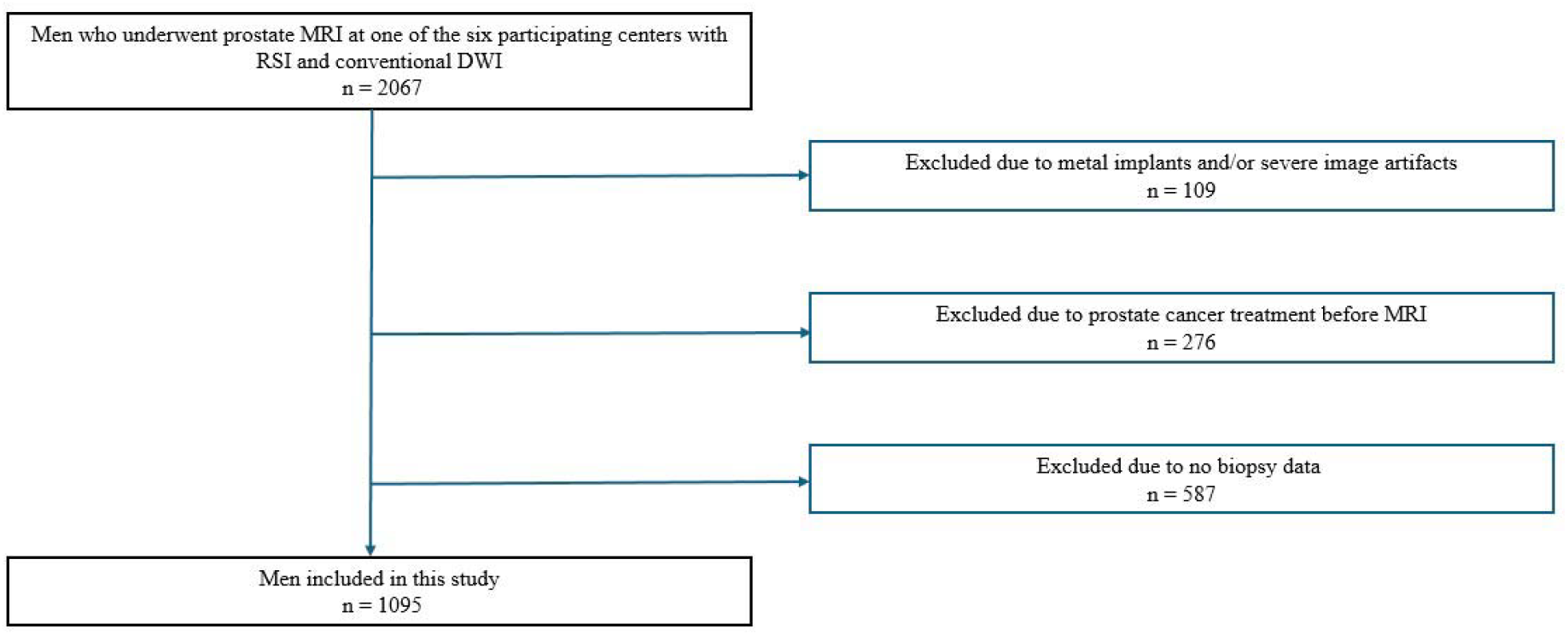
Patient Cohort Flowchart. Flowchart showing how the patient cohort was constructed for this study.

Using conventional DWI data adequate to fit the 4-compartment model (≥4 non-zero *b-*values, n=651 patients), RSIrs_post-hoc_ tended to overestimate RSIrs values (median bias: +156.3; IQR: [49.4, 267.2]; p < 0.001). A similar pattern was observed in the cohort of protocols (n=444) using the 3-compartment model (median: +100.7; IQR: [9.8, 193.8]; p < 0.001) (Figure 2A). The absolute average error was reduced with the RSIrs_combo_ method (median: −58.5; IQR: [−97.6, − 29.0]; p < 0.001) (Figure 2B). Analysis of subgroups based on center and acquisition protocol revealed that this improvement was driven mostly by datasets lacking a high *b*-value in the conventional DWI data. E.g., in data from UCSF (conventional DWI *b*-values: 0, 600, 1000 s/mm^2^), RSIrs_post-hoc_ demonstrated a very large overestimation bias (median: +188.680; IQR: [123.5, 257.9]; p < 0.001), but RSIrs_combo_ greatly reduced the average error (median: −45.6; [−65.8, −13.9]; p < 0.001) (Supplementary Figure 1B). On the other hand, in data from UCSDH (conventional DWI *b*-values: 50, 1000, 1400 s/mm^2^) RSIrs_post-hoc_ showed no significant systematic bias from the reference (median: +3.84; IQR: [−33.1, 48.4]; p=0.445), and RSIrs_combo_ modestly worsened the average estimate (median: −44.528; IQR: [−81.5, −14.4]; p < 0.001) (Supplementary Figure 1D).

**Figure 2:**
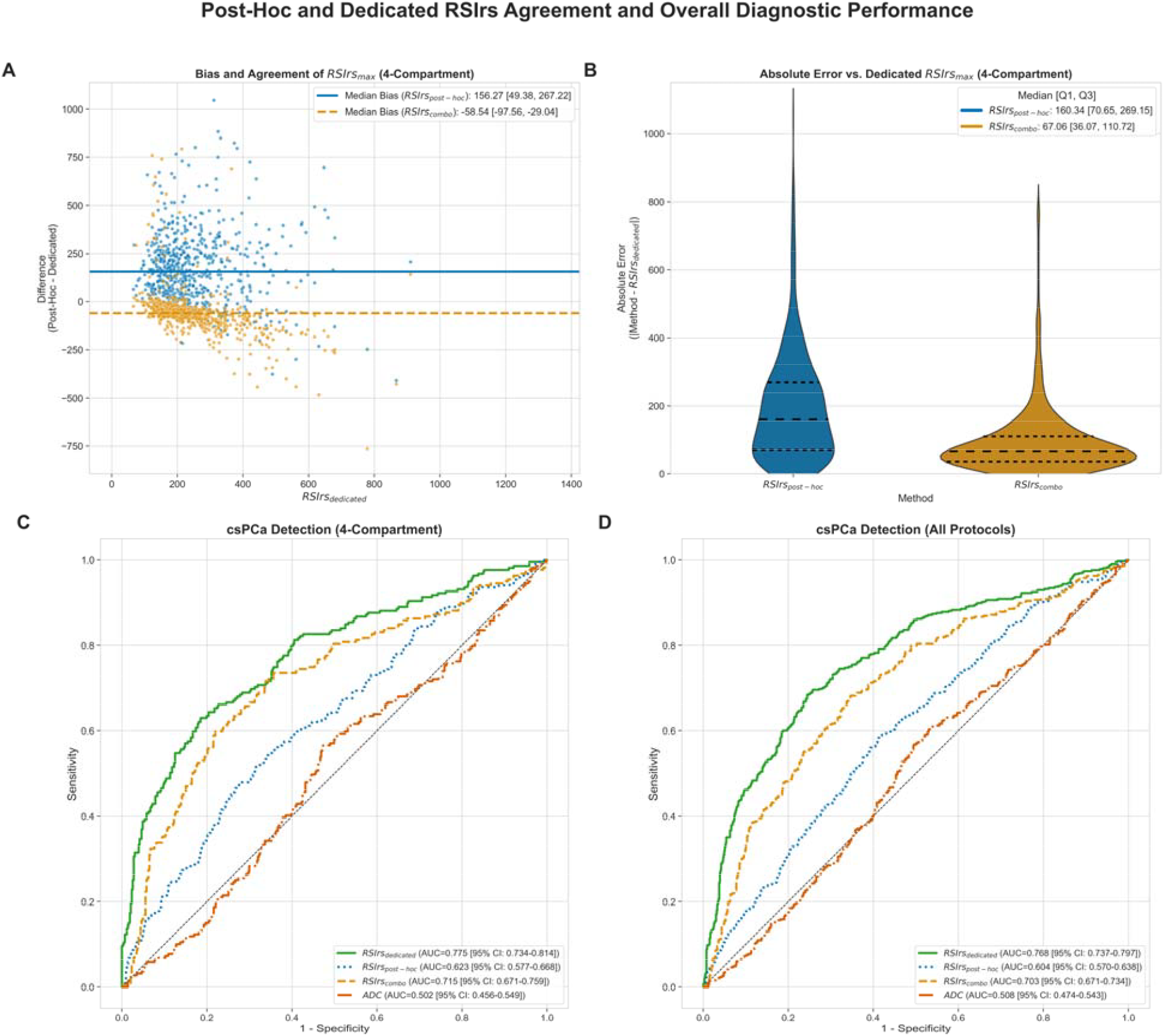
Quantitative Comparison of *Post-Hoc* RSI to Dedicated RSI. Plots A and B show Bland-Altman and violin plots showing the bias, agreement, and absolute error between *post-hoc* RSI methods and dedicated RSI using the 4-compartment RSI model, respectively. Each dot represents an individual patient and their bias of *post-hoc* to dedicated RSIrs; each patient has two dots, one for each *post-hoc* RSIrs. Plots C and D display receiver operating characteristic curves for the detection of clinically significant prostate cancer (csPCa) for 4-compartment model and all data (a 3-compartment RSI model was used if only three non-zero *b*-values were available) of *post-hoc* RSI, dedicated RSI, and the apparent diffusion coefficient (ADC). The plots showcase the hierarchy of performance: dedicated RSI > combination of conventional diffusion weighted imaging (DWI) and RSI > conventional DWI alone > ADC.

Lesion-level evaluation of *post-hoc* RSIrs estimates was feasible in a subset of patients with contoured PI-RADS lesions and corresponding pathology diagnosis from whole-mount histopathology (Figure 3). In PI-RADS defined lesions (n=58), RSIrs_post-hoc_ bias was (median: +179.8 IQR: [64.3, 252.3]; p < 0.001) and RSIrs_combo_ bias was (median: −72.1; IQR: [−113.2, − 24.3]; p < 0.001). In tumors confirmed on whole-mount histopathology (n=64), RSIrs_post-hoc_ bias was (median: −27.6; IQR: [−82.4, 13.3]; p = 0.036), while RSIrs_combo_ bias was (median: −32.5; IQR: [−83.4, −3.0]; p < 0.001).

**Figure 3:**
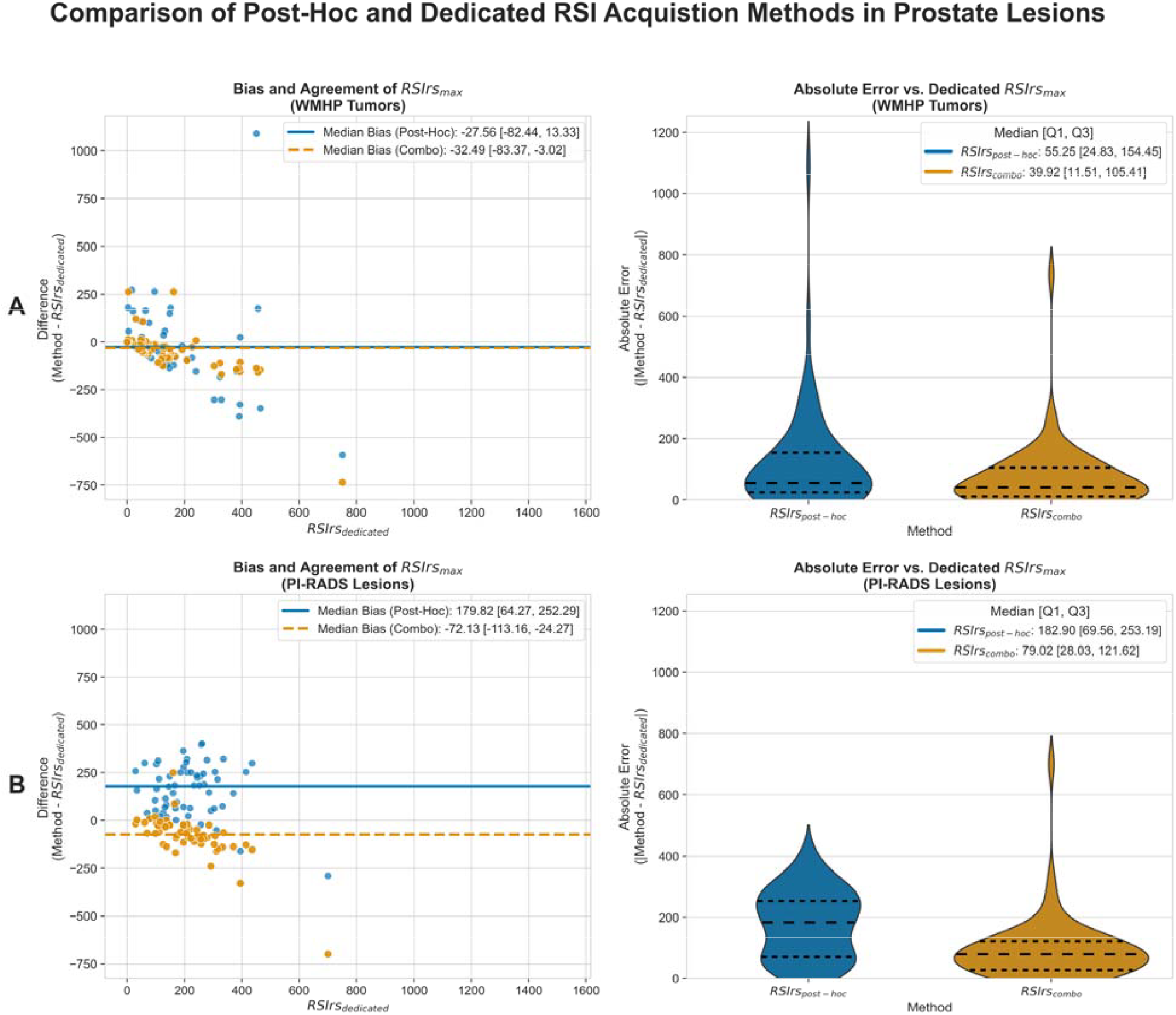
Quantitative Comparison of *Post-Hoc* RSI to Dedicated RSI in Expert-defined Tumors/Lesions. Plots A and B showcase the bias, agreement, and absolute error between *post-hoc* and dedicated RSI metrics within prostate tumors/lesions contoured by clinical experts. In both plots, the left graph shows a bland-altman analysis, and the right graph shows two violin plots comparing RSIrs_post-hoc_ and RSIrs_combo_ to RSIrs_dedicated_. Each dot represents an individual patient and their bias of *post-hoc* to dedicated RSIrs; each patient has two dots, one for each *post-hoc* RSIrs. Agreement with RSIrs_dedicated_ was high for *post-hoc* methods in WMHP tumors, but there is a noticeable discrepancy in PI-RADS lesions, overall (which may or may not be cancer). RSIrs_combo_ improves agreement in both patient subsets overall.

Quantitative csPCa detection was significantly better with dedicated RSI acquisition than with *post-hoc* methods, though all RSIrs estimates had better performance than ADC (Figure 2D). RSIrs_dedicated_ achieved the highest accuracy (AUC: 0.77; 95% CI: [0.74,0.80]). RSIrs_combo_ performed second best (AUC: 0.70; 95% CI: [0.67,0.74]), RSIrs_post-hoc_ third (AUC: 0.60; 95% CI: [0.57,0.64]) and ADC last (AUC: 0.51; 95% CI: [0.47,0.54]).

We repeated the primary analyses with a 3-compartment model for those subsets of data where the conventional DWI included insufficient *b*-values to permit fitting a 4-compartment model. The patterns of results were similar to those in the 4-compartment model analyses (see Supplementary Figure 2).

## Discussion

In data from six imaging centers, we found that *post-hoc* estimation of RSIrs is feasible and that *post-hoc* RSIrs is still superior to ADC as a quantitative metric for prostate/patient-level automated csPCa detection. *Post-hoc* RSIrs can be used in retrospective analysis of datasets that did not have a dedicated RSI acquisition or as a temporary solution during the early stages of implementing dedicated RSI. However, *post-hoc* RSI methods are not a complete substitute for a dedicated RSI acquisition. Estimation of RSIrs is best when a high *b*-value (>1000 s/mm^2^) is acquired. These results can also inform efforts to balance constraints on acquisition protocols.

Where feasible, a dedicated RSI acquisition is advised to optimize quantitative results. Given that ADC and average *b-*value maps can be calculated from the DWI data in a multi-*b-*value RSI acquisition, if only one DWI acquisition can be performed, a dedicated RSI protocol can yield both conventional and *post-hoc* RSIrs maps and metrics. If a dedicated RSI acquisition is not feasible, RSIrs may still be estimated.

While the quantitative performance of *post-hoc* RSI was superior to ADC, it is worth noting that the *post-hoc* approach introduces systematic bias/error in the quantitative values. Clinically relevant thresholds like those described for dedicated RSIrs [Rojo Domingo et al. 2024] likely need to be re-calibrated for *post-hoc* RSIrs (Figure 4). This calibration would need to be performed for each acquisition protocol, as conventional DWI protocols can vary markedly across centers. Conversely, previous work has shown dedicated RSIrs is relatively robust to variation in center, protocol, scanner, and patient demographics [Do et al. 2024].

**Figure 4:**
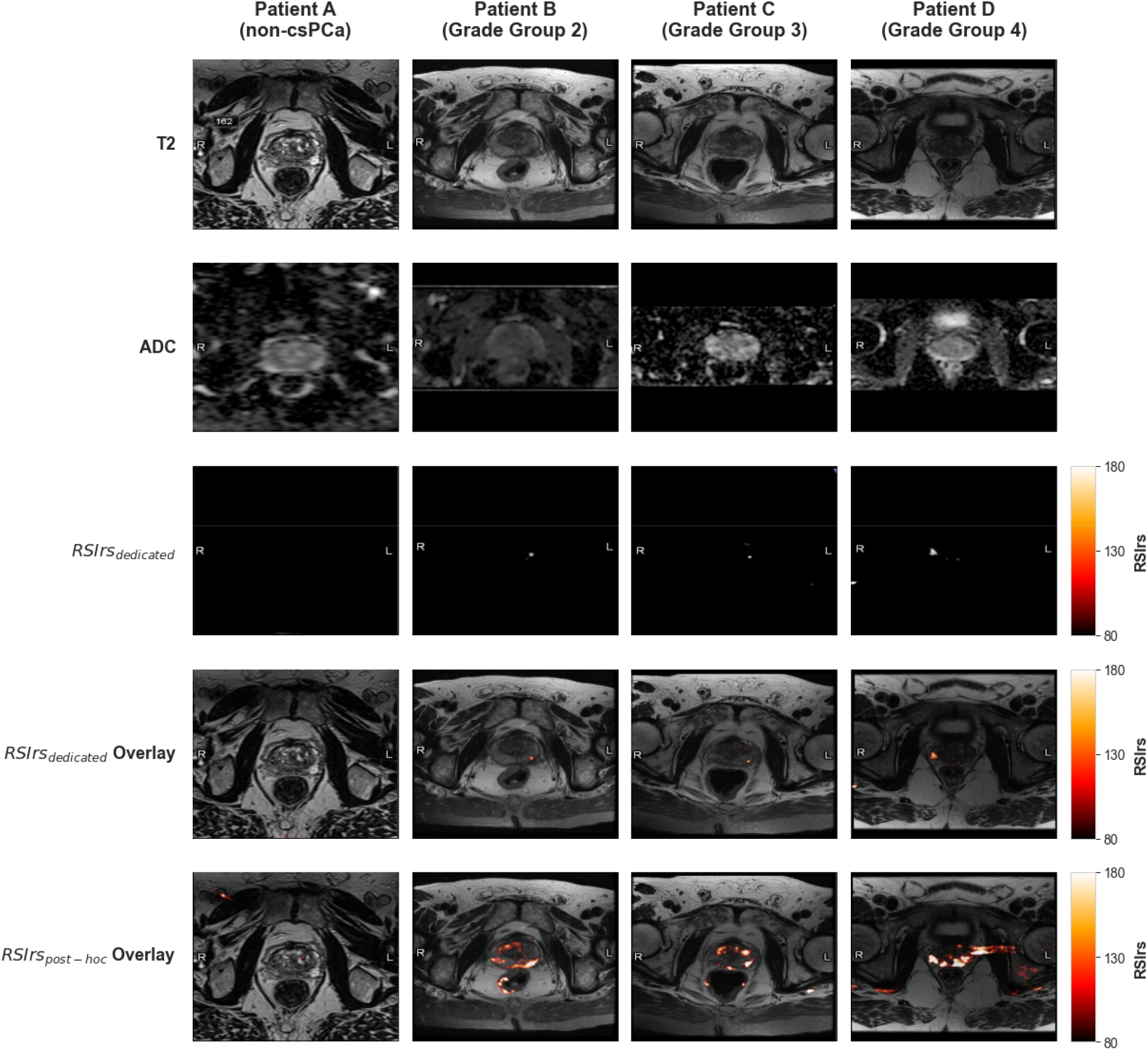
Examples of Quantitative Biomarker Maps. Four representative patients (A-D). Patient A had no csPCa while patients B-D had increasingly aggressive csPCa (B: Grade Group (GG) 2, C: GG3, D: GG4) and underwent radical prostatectomy. The corresponding slice on MRI is displayed in subsequent rows *T*_*2*_*-*weighted imaging, apparent diffusion coefficient (ADC), dedicated RSIrs, and RSIrs (two versions) overlaid on *T*_*2*_. The color bar indicates the signal intensity windows used for all RSIrs maps. All patients had a 4-compartment *post-hoc* RSIrs; a high b-value was available from conventional DWI data for Patients B, and C. Overall, RSIrs_dedicated_ has the greatest agreement with location of csPCa tissue on RSI.

The strong csPCa detection performance of RSIrs_combo_ is encouraging. It demonstrates that the performance of *post-hoc* RSI methods can be substantially improved by incorporating even just one high *b*-value into the standard clinical protocol. Overall, this study provides two alternative strategies to retrospectively analyze conventional DWI data and assist centers who are hesitant to alter their standard protocols to implement RSI, by applying *post-hoc* RSI accordingly based on the availability of high *b*-value data. A simple addition of one high *b*-value acquisition (i.e., *b* = 1500 s/mm^2^) would only minimally increase protocol acquisition time to gain the benefits of RSIrs.

Consistent with prior studies, whole-gland minimum ADC is a poor marker for presence of csPCa [Zhong et al. 2023, Rojo Domingo et al. 2025]. It is known that low ADC values are not cancer-specific and are commonly found in benign prostatic tissue, including the anterior fibromuscular stroma and BPH nodules. Furthermore, a single minimum voxel value is highly sensitive to image noise. We reiterate that minimum ADC within the prostate is not how ADC is routinely used in clinical practice, where an experienced radiologist first identifies a suspicious lesion and then semi-quantitatively evaluates the focal ADC values (e.g., mean ADC) of the lesion in comparison to normal-appearing prostate tissue. An advantage of RSIrs is its potential as quantitative biomarker that has utility without first identifying a lesion (a process that is subject to inter-reader variability and is dependent on reader expertise). ADC is used quantitatively in the present study for direct comparison to RSIrs. We found that even *post-hoc* RSIrs significantly distinguishes prostates that contain csPCa from those that do not, illustrating the value of parsing signal from hypercellular, cancerous tissue from other causes of diminished diffusion.

A limitation of this study is its reliance on PI-RADS interpretation and biopsy results as the ground truth where interpretation heavily relies on reader expertise. Biopsy techniques are also prone to error. On the other hand, whole-mount histopathology is only available for patients who are first diagnosed with csPCa, are candidates for surgery, and who elect to undergo radical prostatectomy. Thus, biopsy results, despite their imperfection, are the relevant clinical gold standard. This study is also limited by a lack of central radiology or pathology review. For some protocols, we note that a *b* = 0 acquisition was not available, so the lowest available *b*-value was used to estimate RSIrs. Finally, even data from seven centers will not represent all possible conventional DWI protocols.

## Conclusion

RSIrs estimated *post-hoc* from conventional DWI offers a significant improvement over standard ADC as a patient-level quantitative biomarker of csPCa. RSIrs calculated from a dedicated, multi-*b*-value acquisition performs best. *Post-hoc* RSIrs can be used in retrospective analyses where dedicated RSI data is not available and could also be implemented clinically at centers with heavy constraints on modifying their DWI protocols.

## Data Availability

All data produced in the present study are available upon reasonable request to the authors

