## Supplementary Material for "Post-hoc Estimation of a Quantitative Restriction Spectrum Imaging Biomarker for Prostate Cancer Detection Using Conventional MRI"

### Supplementary Table 1. MRI Acquisition Parameters by Institution

| **Institution** | **Parameter** | **RSI** | **Conventional DWI (Full-FOV)** | **Conventional DWI (FOCUS)** | **T2-weighted** |
| --- | --- | --- | --- | --- | --- |
| **UCSD CTIPM** | Pulse sequence | Diffusion-weighted EPI | EP/SE | EP/SE | FSE |
|  | TR (ms) | 4500 | 6400 | 4000 | 7000 |
|  | TE (ms) | 76 | 52 | 65 | 100 |
|  | FOV (mm) | 200 x 100 | 360 x 360 | 240 x 144 | 240 x 240 |
|  | Matrix [resampled] | 80 x 48 [128 x 128] | 80 x 128 [256 x 256] | 160 x 96 [256 x 256] | 320 x 320 [512 x 512] |
|  | Slices | 32 | 168 | 32 | 32 |
|  | Slice Thickness (mm) | 3 | 5 | 5 | 3 |
|  | b-values (s/mm²) [samples] | 0[1], 50[6], 800[6], 1500[12], 3000[18] | 0, 50, 1000 | 0, 400 | N/A |
|  | Field Strength (T) | 3 | 3 | 3 | 3 |
| **UCSDH** | Pulse sequence | Diffusion-weighted EPI | EP/SE | EP/SE | FSE |
|  | TR (ms) | 4000 | 5000 | 5000 | 5300 |
|  | TE (ms) | 69 | 60 | 57 | 100 |
|  | FOV (mm) | 240 x 120 | 280 x 280 | 240 x 144 | 200 x 200 |
|  | Matrix [resampled] | 96 x 48 [256 x 256] | 128 x 128 [256 x 256] | 160 x 96 [256 x 256] | 320 x 320 [512 x 512] |
|  | Slices | 16 | 52 | 30 | 32 |
|  | Slice Thickness (mm) | 6 | 5 | 4 | 3 |
|  | b-values (s/mm²) [samples] | 0[1], 500[8], 1000[8], 2000[16] | 0, 50, 1000 | 0, 1400 | N/A |
|  | Field Strength (T) | 3 | 3 | 3 | 3 |
| **URMC** | Pulse sequence | Diffusion-weighted EPI | EP | - | FSE |
|  | TR (ms) | 3800 | 4900 | - | 4800 |
|  | TE (ms) | 85 | 89 | - | 104 |
|  | FOV (mm) | 52 x 52 | 250 x 250 | - | 180 x 180 |
|  | Matrix [resampled] | 52 x 100 [104 x 200] | 114 x 114 [114 x 144] | - | 384 x 365 [384 x 384] |
|  | Slices | 22 | 23 | - | 32 |
|  | Slice Thickness (mm) | 4 | 3.5 | - | 3 |
|  | b-values (s/mm²) [samples] | 0[1], 500[6], 1000[6], 2000[6] | 50, 400, 800 | - | N/A |
|  | Field Strength (T) | 3 | 3 | - | 3 |
| **MGH** | Pulse sequence | Diffusion-weighted EPI | EP/SE | EP/SE | FSE |
|  | TR (ms) | 4500 | 4600 | 2000 | 3937 |
|  | TE (ms) | 59 | 54 | 60 | 169 |
|  | FOV (mm) | 240 x 120 | 320 x 320 | 200 x 120 | 160 x 160 |
|  | Matrix [resampled] | 96 x 48 [128x128] | 128 x 128 [256 x 256] | 132 x 80 [256 x 256] | 360 x 224 [1024 x 1024] |
|  | Slices | 16 | 45 | 26 | 40 |
|  | Slice Thickness (mm) | 6 | 3 | 4 | 3 |
|  | b-values (s/mm²) [samples] | 0[1], 500[6], 1000[6], 2000[12] | 50, 800 | 200, 2000 | N/A |
|  | Field Strength (T) | 3 | 3 | 3 | 3 |
| **UTHSCA** | Pulse sequence | Diffusion-weighted EPI | - | EP | FSE |
|  | TR (ms) | 5500 | - | 4600 | 4710 |
|  | TE (ms) | 98 | - | 80 | 100 |
|  | FOV (mm) | 300 x 155 | - | 260 x 180 | 180 x 180 |
|  | Matrix [resampled] | 128 x 66 [108 x 128] | - | 160 x 110 [256 x 256] | 240 x 320 [320 x 320] |
|  | Slices | 25 | - | 25 | 30 |
|  | Slice Thickness (mm) | 3 | - | 3 | 3 |
|  | b-values (s/mm²) [samples] | 0[1], 600[30], 1200[30], 1800[30] | - | 0[1], 400[18], 800[18], 1000[18] | N/A |
|  | Field Strength (T) | 3 | - | 3 | 3 |
| **UCSF** | Pulse sequence | Diffusion-weighted EPI | - | EP/SE | FSE |
|  | TR (ms) | 4500 | - | 4800 | 2964 |
|  | TE (ms) | 73 | - | 46 | 150 |
|  | FOV (mm) | 200 x 200 | - | 200 x 100 | 220 x 220 |
|  | Matrix [resampled] | 256 x 256 [256 x 256] | - | 128 x 64 [256 x 256] | 512 x 512 [320 x 320] |
|  | Slices | 35 | - | 37 | 36 |
|  | Slice Thickness (mm) | 3 | - | 3 | 3 |
|  | b-values (s/mm²) [samples] | 0[5], 100[6], 800[12], 1400[12], 2500[18] | - | 0[1], 600[4], 1000[4] | N/A |
|  | Field Strength (T) | 3 | - | 3 | 3 |

**Scanner Information**

| **Cohort** | **Scanner Models** | **Number of Stations** |
| --- | --- | --- |
| **UCSD CTIPM** | GE Healthcare Discovery MR750, GE Healthcare Signa Premier | 4 |
| **UCSDH** | GE Healthcare Discovery MR750, GE Healthcare Signa Premier | 4 |
| **URMC** | SIEMENS Skyra | 2 |
| **MGH** | GE Healthcare Signa Premier | 1 |
| **UTHSCA** | SIEMENS Skyra | 3 |
| **UCSF** | GE Healthcare Signa Premier | 2 |
| **Total** |  | **16** |

*Abbreviations: CTIPM, Center for Translational Imaging and Precision Medicine; DWI, diffusion-weighted imaging; EPI, echo-planar imaging; EP/SE, echo-planar/spin-echo; FOV, field of view; FSE, fast spin echo; MGH, Massachusetts General Hospital; RSI, restriction spectrum imaging; TE, echo time; TR, repetition time; UCSD(H), University of California San Diego (Health); UCSF, University of California San Francisco; URMC, University of Rochester Medical Center; UTHSCA, University of Texas Health Science Center at San Antonio*

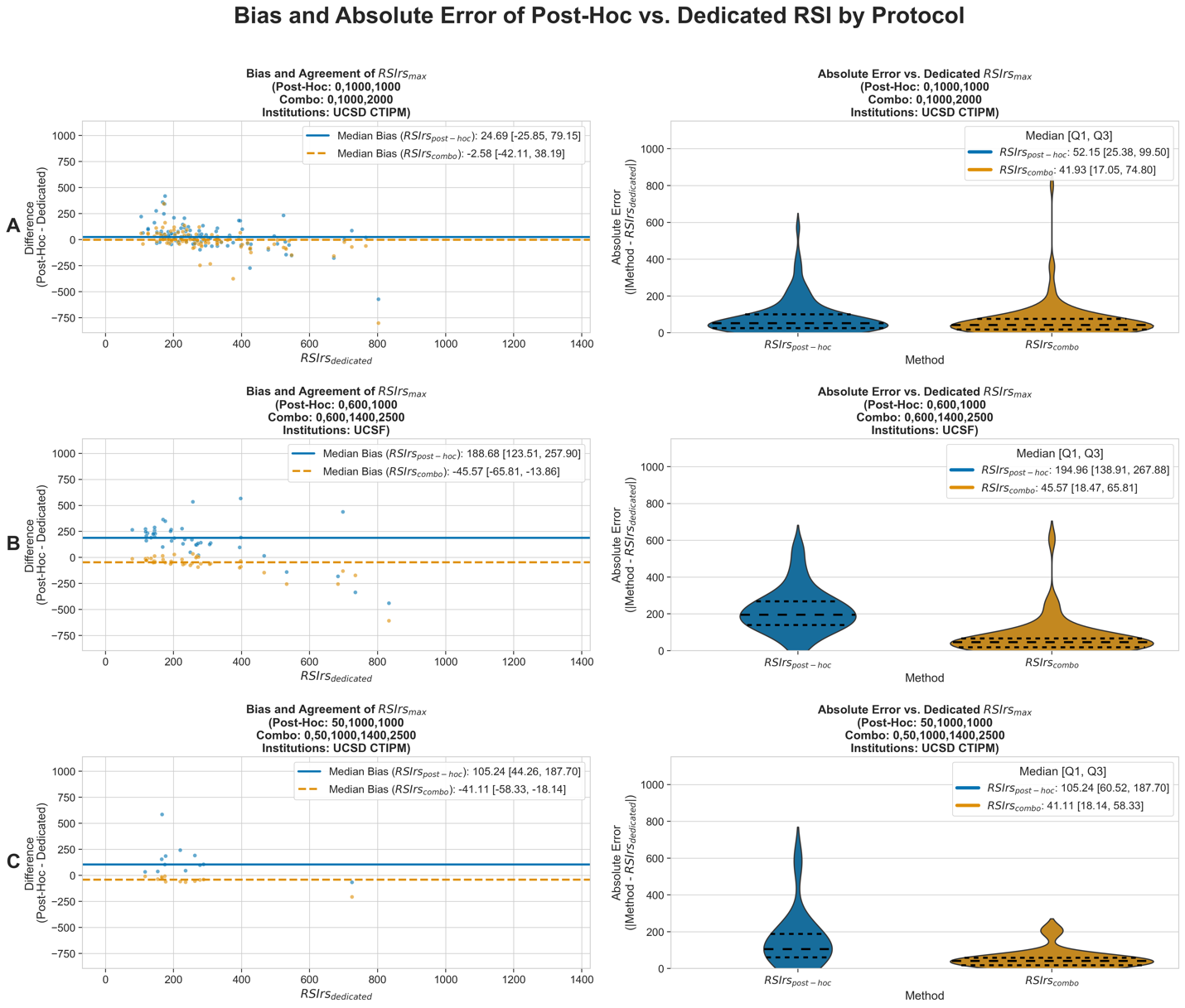

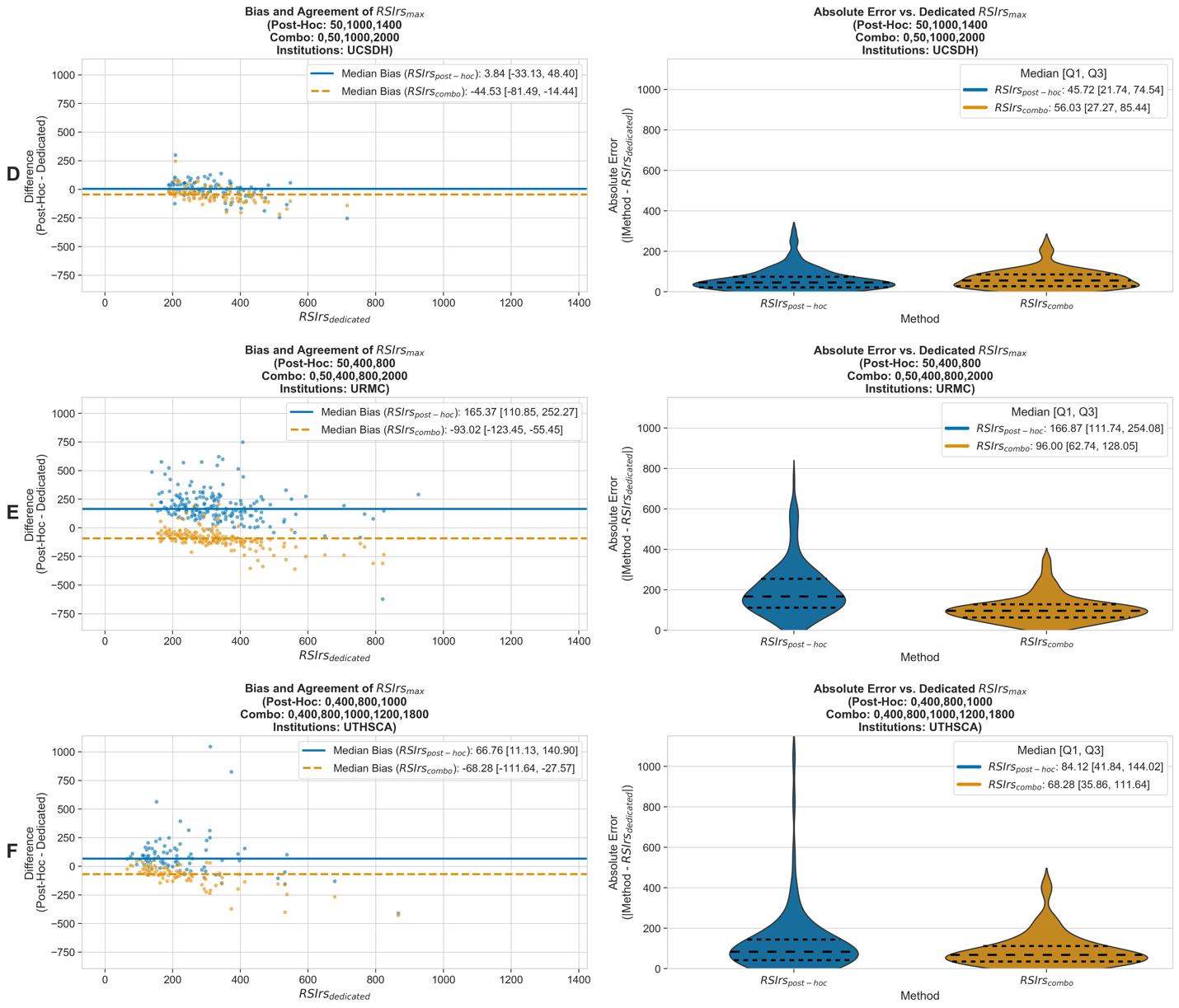

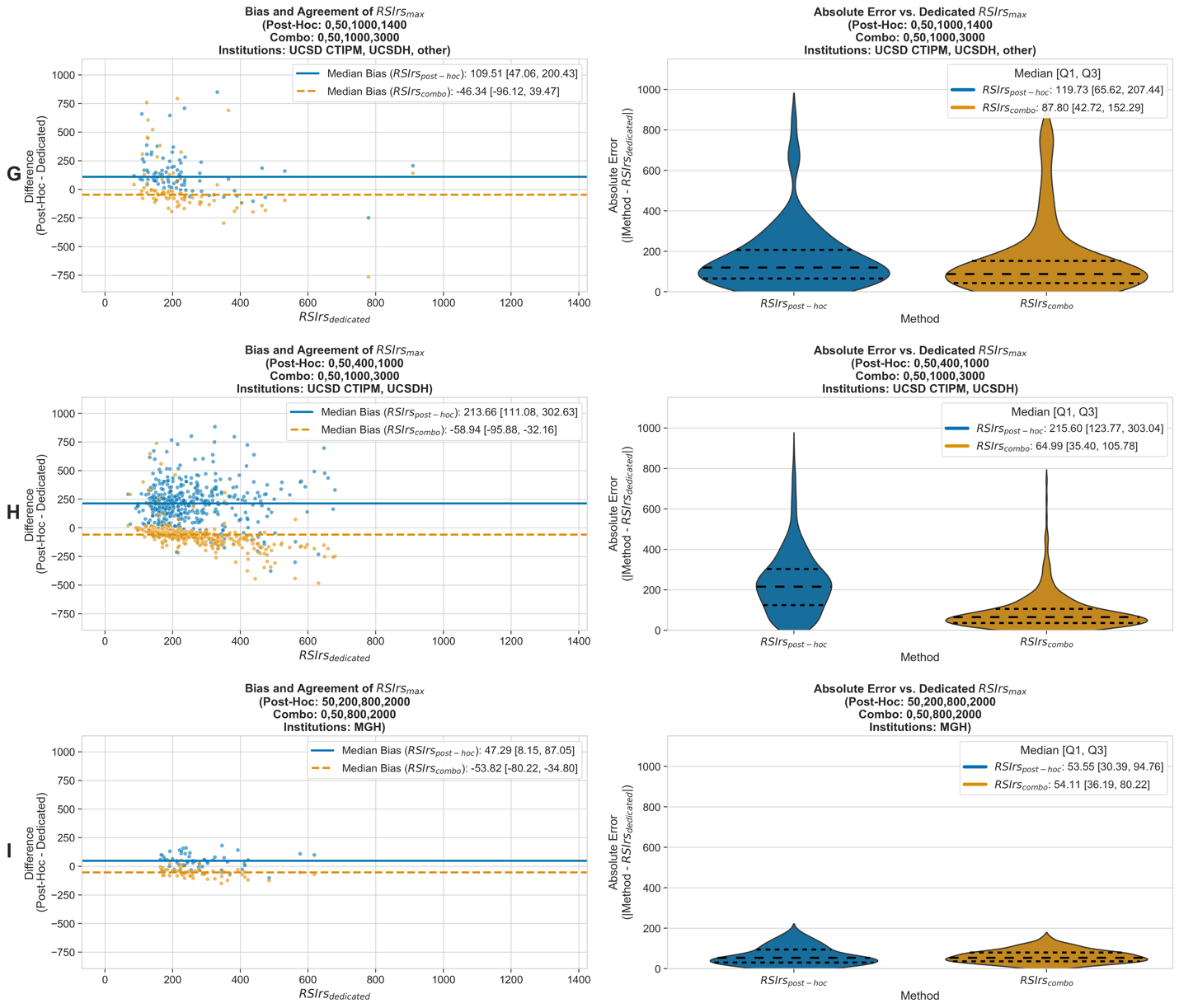

**Supplementary Figure 1: Comparison of *Post-hoc* to dedicated RSI in Prostate Lesions**

Plots A-I showcase the bias, agreement, and absolute error of *post-hoc* versus dedicated RSI for each of the nine individual acquisition protocols evaluated in this study. For each protocol, a Bland-Altman plot on the left visualizes the systematic bias and limits of agreement between the two *post-hoc* methods and dedicated RSI. On the right, two violin plots illustrate the distribution and median of the absolute error, showcasing how RSIrs_combo_ reduces the error in RSIrs estimation compared to RSIrs_post-hoc_. However, it should be noted that this improvement only appears to occur when the conventional DWI data did not already have a high *b*-value. Between the protocols that did (D, I and G), the protocols in D and I had worse agreement between RSIrs_combo_ and RSIrs_dedicated_, although the protocol in G did not.

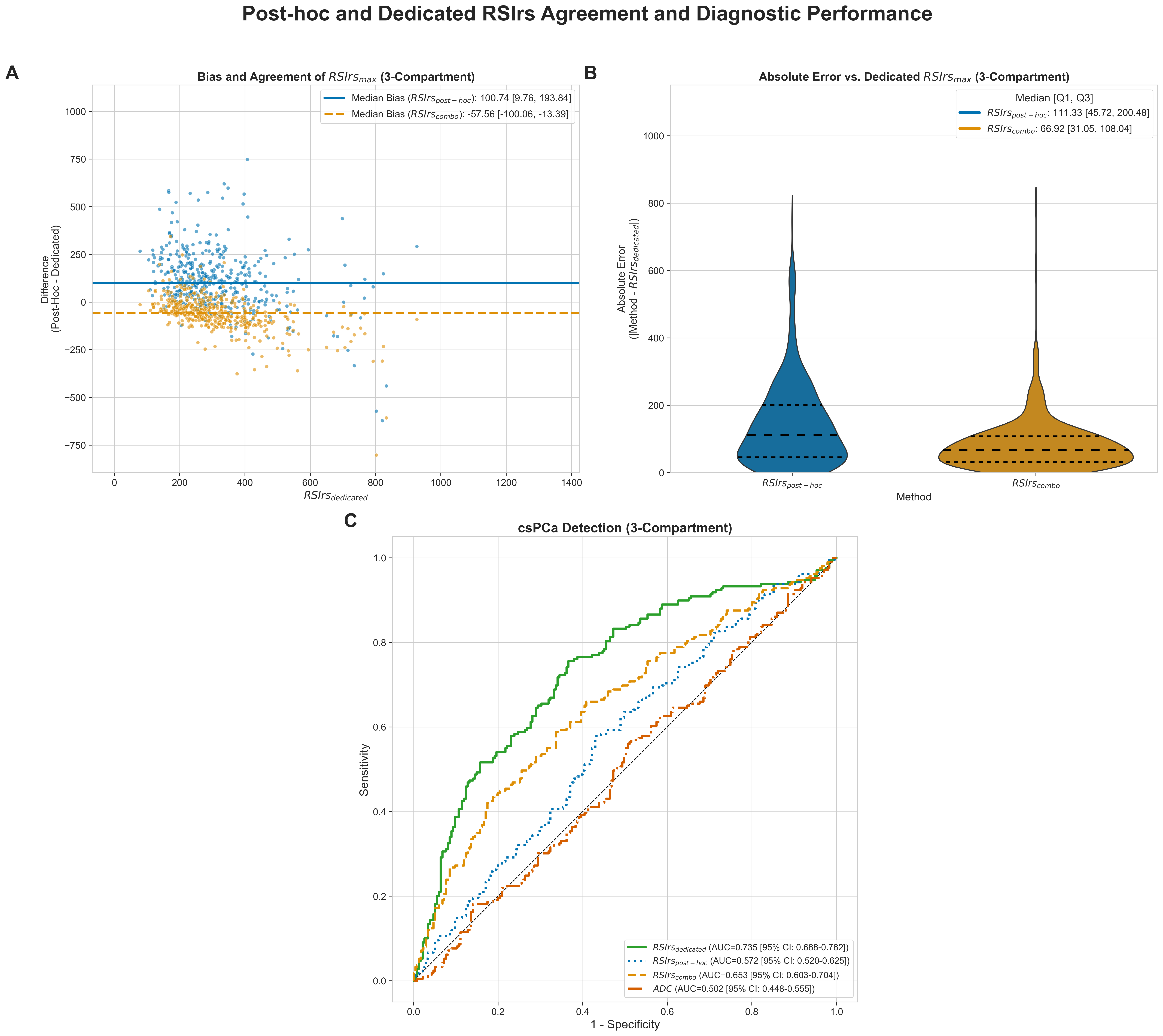

**Supplementary Figure 2: Quantitative Comparison of *Post-Hoc* to Dedicated RSI for 3-Compartment Model**

Plots A and B show Bland-Altman and violin plots comparing the bias, agreement, and absolute error between *post-hoc* RSI methods and dedicated RSI using data compatible with the 3-compartment RSI model. Plot C display Area under the corresponding receiver operating characteristic (AUC) curve for the detection of clinically significant prostate cancer (csPCa) for *post-hoc* RSIrs, dedicated RSIrs, and the apparent diffusion coefficient (ADC). The plots showcase the hierarchy of performance dedicated RSI > combination of conventional diffusion weighted imaging (DWI) and RSI > conventional DWI alone > ADC.
